# Subcortical Functional Connectivity Gradients in Temporal Lobe Epilepsy

**DOI:** 10.1101/2023.01.08.23284313

**Authors:** Alfredo Lucas, Sofia Mouchtaris, Eli J. Cornblath, Nishant Sinha, Lorenzo Caciagli, Peter Hadar, James J. Gugger, Sandhitsu Das, Joel M. Stein, Kathryn A. Davis

## Abstract

**Background and Motivation:** Functional gradients have been used to study differences in connectivity between healthy and diseased brain states, however this work has largely focused on the cortex. Because the subcortex plays a key role in seizure initiation in temporal lobe epilepsy (TLE), subcortical functional-connectivity gradients may help further elucidate differences between healthy brains and TLE, as well as differences between left (L)-TLE and right (R)-TLE.

**Methods:** In this work, we calculated subcortical functional-connectivity gradients (SFGs) from resting-state functional MRI (rs-fMRI) by measuring the similarity in connectivity profiles of subcortical voxels to cortical gray matter voxels. We performed this analysis in 23 R-TLE patients and 32 L-TLE patients (who were otherwise matched for age, gender, disease specific characteristics, and other clinical variables), and 16 controls. To measure differences in SFGs between L-TLE and R-TLE, we quantified deviations in the average functional gradient distributions, as well as their variance, across subcortical structures.

**Results:** We found an expansion, measured by increased variance, in the principal SFG of TLE relative to controls. When comparing the gradient across subcortical structures between L-TLE and R-TLE, we found that abnormalities in the ipsilateral hippocampal gradient distributions were significantly different between L-TLE and R-TLE.

**Conclusion:** Our results suggest that expansion of the SFG is characteristic of TLE. Subcortical functional gradient differences exist between left and right TLE and are driven by connectivity changes in the hippocampus ipsilateral to the seizure onset zone.

## Introduction

Epilepsy is a brain disorder characterized by recurrent seizures that result from abnormal synchronization of neural activity. While seizures are well controlled with anti-seizure medications in many individuals with epilepsy, nearly a third of them are pharmacoresistant^1^. Many of these cases occur in patients with temporal lobe epilepsy (TLE), the most common type of focal epilepsy^2^. In these patients, seizures originate in the mesial temporal lobe (hippocampus, parahippocampal gyrus, and amygdala) or in the temporal neocortex. In unilateral TLE, seizures can be localized to either the left (L-TLE) or the right temporal lobe (R-TLE). Initially, L-TLE and R-TLE were thought to be symmetric disorders, but recent work has provided evidence to the contrary ^3–6^. However, current literature disagrees as to whether L- or R-TLE has more extensive abnormalities^7^.

Epilepsy is being increasingly conceptualized as a network disorder, with abnormal connections across the brain^8,9^. These abnormalities can be quantified and compared across patients via the application of graph-theoretical approaches to measures of functional connectivity, generated from functional MRI (fMRI)^10–12^. In TLE, these approaches have demonstrated both local and global epileptic network abnormalities despite focal seizure localization^13–15^.

Functional connectivity matrices can additionally be used to generate functional connectivity gradients, using linear and non-linear dimensionality reduction techniques^16^. Functional connectivity gradients provide a simple, yet efficient, representation of connectivity across the brain, where regions close to each other in gradient space have similar connectivity profiles^17^. Initial work demonstrated that the first principal cortical gradient anchors unimodal sensory and motor regions at one end, and transmodal regions involved in higher level processing, such as those belonging to frontoparietal and default mode networks at the opposite end. In addition, the second principal gradient can differentiate between visual and somatosensory/motor regions^18^. These functional gradients are altered in disease states^19–21^. In idiopathic generalized epilepsy, the values taken by the first gradient are expanded, suggestive of increased differentiation in connectivity profiles compared to controls^20^. In TLE, a similar functional connectivity distance metric (distinct from gradients) was also found to be contracted in temporoinsular and prefrontal networks relative to controls^22^. Functional activation derived from task-based fMRI compared to resting state functional gradients probe functional reorganization due to cognitive impairment in epilepsy^13,23^. In subcortical-to-cortical functional connectivity gradients, subcortical voxels/regions with similar connections to cortical gray matter regions will be close together in gradient space. These gradients were used to elucidate the topographical organization of the subcortex^24^, and demonstrate a unimodal to transmodal organization in the thalamus similar to that in the cortex^20,25^. To our knowledge, no work has explored the effects of epilepsy on subcortical-to-cortical functional gradients, despite the prominent role of subcortical structures in the pathophysiology of epilepsy.

In this study, we use subcortical functional connectivity gradients generated from 3T resting-state fMRI to compare TLE to healthy controls, and to investigate patterns of reorganization that may be specific to L-TLE and R-TLE. We tested whether the first principal gradient was expanded in TLE as compared to healthy controls, in line with prior work^13,20^. We also tested whether gradient distributions in key subcortical structures that contribute to seizures were altered in patients with TLE. Lastly, we explored whether changes seen in L-TLE differed from those seen in R-TLE, which would further contribute to the evidence in favor of key functional connectome differences between the two disease states.

## Methods

### Patient Demographics

Data acquisition for this study was approved by the institutional review board of the University of Pennsylvania. A total of 55 temporal lobe epilepsy (TLE) patients were included in this study. Localization of seizure focus was determined during the Penn Epilepsy Surgical Conference (PESC) following evaluation of various clinical, neuroimaging, and neurophysiological data including: seizure semiology, neuropsychological testing, MRI, positron emission tomography (PET), scalp EEG, and intracranial EEG findings. Thirty-one patients had left-sided seizure onset zone (SOZ) lateralization (L-TLE), and 24 patients had right-sided SOZ lateralization (R-TLE). Age, gender, disease duration, MRI lesional status, and history of focal to bilateral tonic-clonic seizures (BTCS) are reported in **Table 1**. No statistically significant differences between demographic variables were found between the L-TLE and R-TLE groups. Sixteen age and gender matched controls (mean age 32±11) were also included in the study.

**Table 1.**
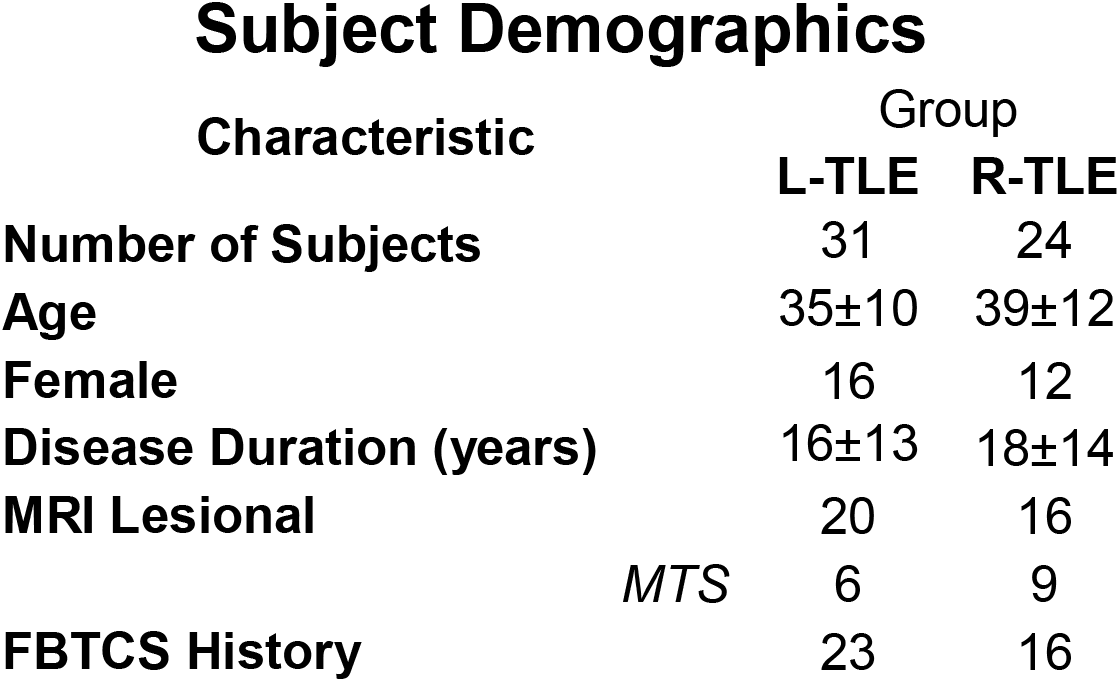
Subject Demographics. Table demographics showing number of subjects, age, sex, disease duration in years, MRI lesional status, and history of focal to bilateral tonic-clonic seizures (BTCS). MTS: mesial temporal sclerosis.

### Image Acquisition

For 46/55 TLE patients and 8/16 controls, we used a Siemens 3T Magnetom PrismaFit scanner. rs-fMRI data were acquired during a 9-min interval with an axial, 72-slice gradient echo-planar sequence, TE/TR=37/800ms, with a 2mm isotropic voxel size (protocol 1). For the remaining 9/55 TLE patients and 8/16 controls of the Penn cohort, we used a Siemens 3T Magnetom Trio scanner. For this subset of patients, resting-state fMRI data were acquired during a 6-min interval with an axial, 72-slice gradient echo-planar sequence, TE/TR=37/800ms, with a 2mm isotropic voxel size (protocol 2). High-resolution T1-weighted images, with a sagittal, 208-slice MPRAGE sequence, TE/TR=2.24/2400ms, with a 0.8mm isotropic voxel size were acquired in all participants.

### Neuroimaging Processing

We used fMRIPrep^26^ to perform brain extraction and segmentation of individual T1-weighted (T1w) images, registration of rs-fMRI data to individual T1w and MNI template space, and time-series confound estimation. We used the fMRIPrep output data as our input to the xcpEngine post-processing pipeline for 36-parameter confound regression, demeaning, detrending and temporal filtering^27^⍰. Complete details about the functional and anatomical processing pipelines can be found in previous work^28^.

### Subcortical Gradient Calculation

Using the Harvard-Oxford cortical and subcortical atlases, we separated the fMRI time-series data into subcortical voxels [*t* seconds x *a* voxels] and cortical grey matter voxels [*t* seconds x *b* voxels] (Figure 1A,B). In our dataset *a=9,159, b=138,522* and *t=675* for protocol 1 and *t=450* for protocol 2. For computational reasons, and given that there were much fewer timepoints than there were cortical gray matter voxels (*t<<b*), the cortical grey matter time-series data was reduced using principal component analysis (PCA) to a dimensionality of [*t* x *t-1]*, following an approach analogous to Haak et al.^16^ (Figure 1C). We then computed the Pearson correlation of every subcortical voxel’s time-series to each principal component, resulting in a [*a* voxels x *t-1*] matrix containing the functional connectivity of each subcortical voxel to each cortical voxel in principal component space (Figure 1D). This resulting functional connectivity matrix was thresholded with only the top 10% of connections in each row remaining^20^. The remaining steps of the gradient calculations were then completed using the BrainSpace Toolbox ^29^. First, the functional connectivity matrix was transformed into a similarity matrix by taking the Pearson correlation between every pair of rows [*a* voxels x *a* voxels] (Figure 1D). Then, the gradients were generated using diffusion map embedding (Figure 1E). For computational considerations, only the first 250 gradients were generated.

**Figure 1.**
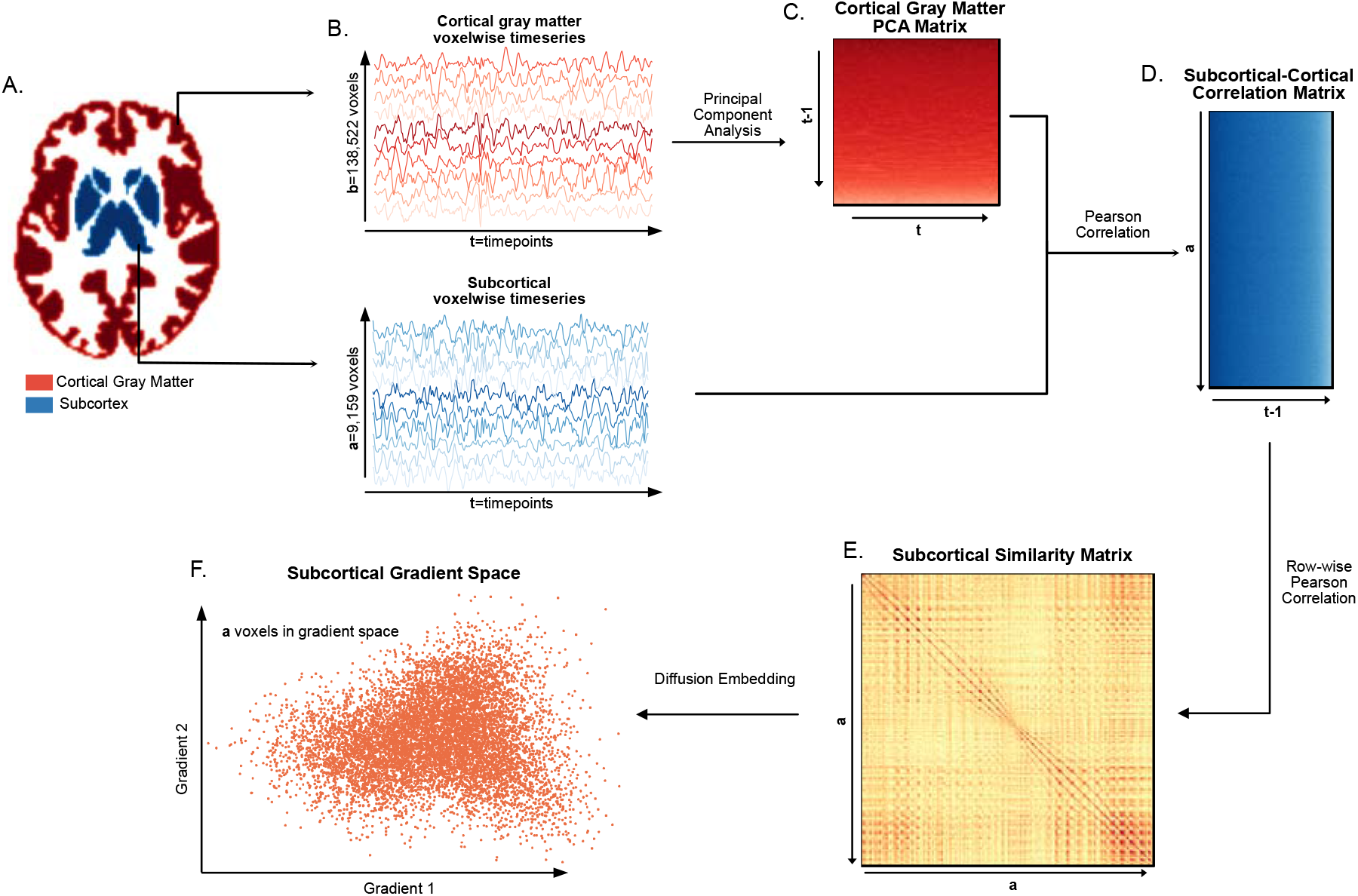
Subcortical Functional Gradient Generation: For each subject, **A**. the subcortical voxels and the cortical gray matter voxels were identified using the Harvard-Oxford subcortical and cortical atlases respectively, and **B**. the timeseries in each of these voxels was extracted. To reduce the computational complexity of the gradient estimation, **C**. the PCA of the cortical gray matter timeseries was taken, resulting in a cortical gray matter PCA matrix. **D**. The Pearson correlation between the cortical gray matter PCA matrix and the subcortical timeseries matrix was computed, resulting in a subcortical-cortical correlation matrix. **E**. The subcortical similarity matrix was estimated by computing the row-wise Pearson correlation of the subcortical-cortical correlation matrix, and **F**. diffusion embedding was applied to the subcortical similarity matrix resulting in the gradient space representation of the subcortical voxels.

Due to differences in gradient ordering and signs, the gradients needed to be aligned to a template before comparison. Following Hong et al.^30^, we horizontally stacked the gradients of all 71 participants (patients + controls) [*a* voxels by 250 * 71]. Using PCA, the first 250 principal components were generated [*a* voxels x 250]. The gradients of all patients were aligned to this template using a Procrustes alignment^31^, which utilizes linear transformations (i.e. translation, scaling, rotation) to align a source to a target.

To account for differences in fMRI collection protocols, the resulting aligned gradients were harmonized using NeuroCombat^32^. This process regressed out differences in gradients due to protocol, but preserved differences due to seizure lateralization and control status.

### Validation Analysis

To assess the stability of the results, we computed the correlation between the gradients obtained in our original approach, with the gradients obtained through different correlation techniques for similarity matrix calculation (the step between Figure 1D and Figure 1E), and different embedding techniques for gradient calculation (the step between Figure 1E and Figure 1F). In our approach proposed above, the similarity matrix was calculated using Pearson’s correlation coefficient, so we also investigated the use of 5 alternative measures: Gaussian kernels, cosine similarity, normalized angle similarity, and Spearman rank order correlations. Additionally, the gradients were originally calculated using diffusion maps, so we considered the use of Laplacian eigenmaps and PCA. This yielded (5×3=) 15 different gradient variants.

To compare the gradients estimated by different methods, we averaged together the aligned and harmonized first and second gradients of all patients for each method, and computed the magnitude of the Pearson correlation between every pair of gradients. The magnitude was used because the gradients within a single method were aligned to each other but not to the gradients of other methods, so the possibility of the gradients having inverted signs had to be accounted for.

### Statistical Analysis

The gradients of all L-TLE and all R-TLE patients were averaged together to yield a representative gradient distribution for each group. Only the first two gradients were considered in subsequent analyses (relative magnitude of eigenvalue 1 = 0.14 ± 0.05, and eigenvalue 2 = 0.09 ± 0.02). In all analyses, unless otherwise specified, we compared regions ipsilateral and contralateral to the SOZ in one group, with the corresponding ipsilateral and contralateral regions in the other group. Since left and right ROIs were flipped as necessary, both L-TLE and R-TLE had an ipsilateral and a contralateral ROI for comparison. With left-to-right flipping, the left sided ROIs in the L-TLE group were always compared with the right sided ROIs in the R-TLE group. To ensure that the measured differences were not driven by inherent asymmetries between the left and right ROIs, we randomly shuffled the group assignments and computed the statistics of interest between the ROIs in the shuffled groups. This procedure was repeated 2000 times, generating a null distribution for each statistic of interest. The p-value (p_PERM_) was determined by the fraction of the null distribution that exceeds the true value in a two-tailed fashion. Additionally, where indicated, a Bonferroni procedure was used to correct for multiple comparisons (p_BON_).

The distribution spanned by principal gradients 1 and 2 was approximately normal and was estimated as a multivariate normal distribution with a [2 × 1] μ and [2 × 2] Σ. We compared the gradients in ipsilateral and contralateral regions between R-TLE and L-TLE using the Bhattacharyya distance, a metric for measuring the similarity of two multivariate normal distributions. To estimate the significance of the Battacharyya distance, we used the same permutation procedure described above.

### Linear Model for Clinical Variables

In addition to studying the effect of disease laterality on the gradient mean and variance, we also measured the effect of clinical variables on these two metrics. To do so, we used a linear model defined as:

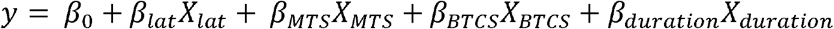

where *y* is either the subject level mean or variance of the gradient, *X*_*lat*_ is an indicator for laterality (L-TLE vs. R-TLE), *X*_*MTS*_ is an indicator variable for the presence of mesial temporal sclerosis (MTS) as determined by structural imaging, *X*_*BTCS*_ is an indicator variable for the presence of a history of bilateral tonic-clonic seizures, and *X*_*duration*_ is a continuous variable that represents the number of years a subject has had epilepsy for. The above model was applied for each subcortical ROI.

## Results

### Subcortical functional connectivity gradients follow a lower to higher computational hierarchy along the first principal gradient

We estimated the functional connectivity signature across subcortical voxels of TLE patients by computing their functional connectivity gradients from subcortical voxels to cortical gray matter voxels. The first two principal gradient values for each subcortical ROI resulted in separate functional clusters, mostly following the anatomical organization of the subcortex (Figure 2, 3A). Interestingly, voxels within functionally related, but anatomically separate regions, such as the caudate and the putamen, had substantial overlap in gradient space. Further dividing each ROI into ipsilateral and contralateral demonstrates that ipsilateral ROIs have similar gradient distributions as their contralateral counterparts (Supplementary Figure 1).

**Figure 2.**
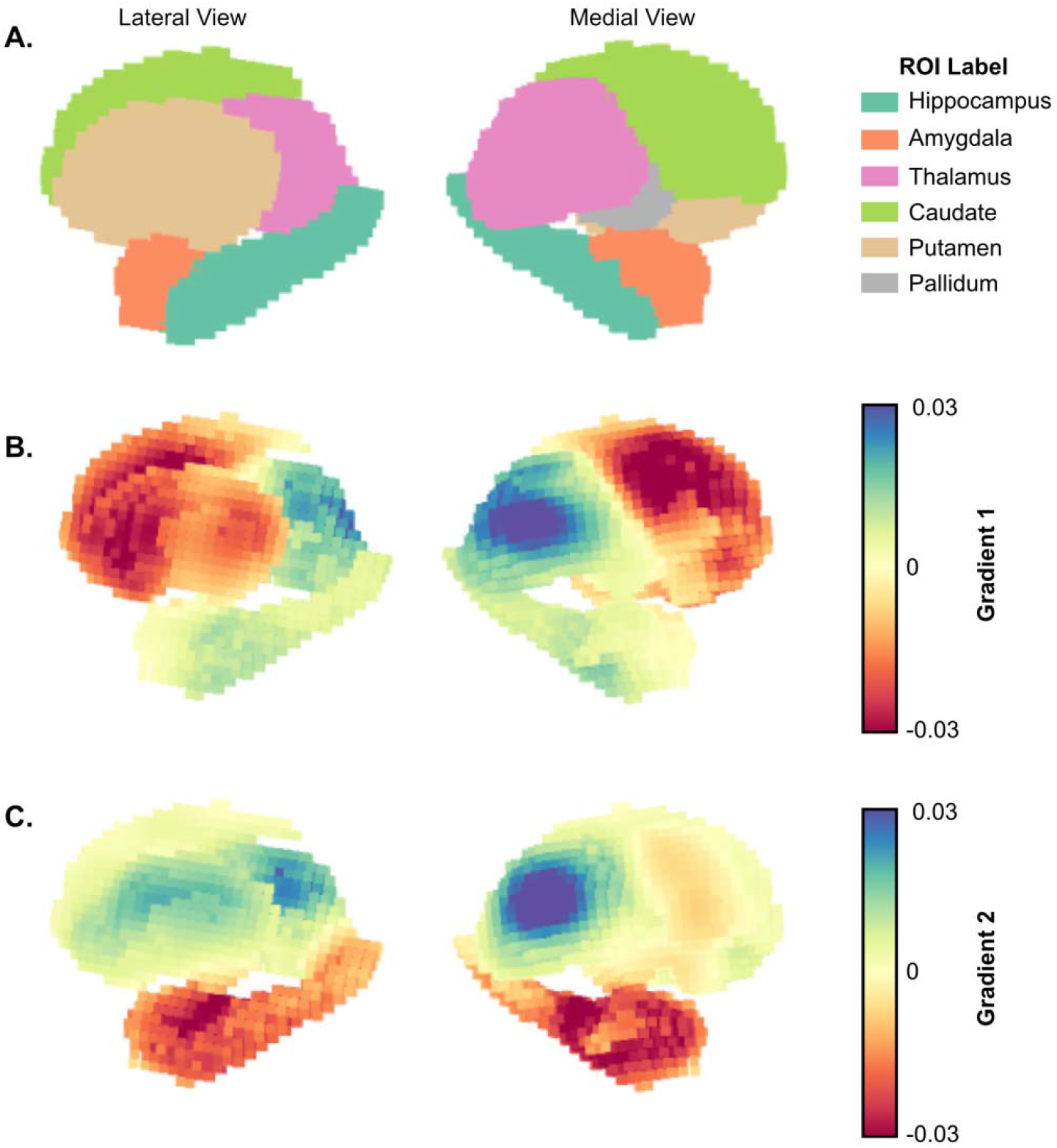
Gradient projection on the subcortical surface: **A**. Lateral (left) and medial (right) view of subcortical regions of interest (ROI) with corresponding labels. Subcortical surface projection of the average across all TLE subjects for **B**. gradient 1 and **C**. gradient 2.

**Figure 3.**
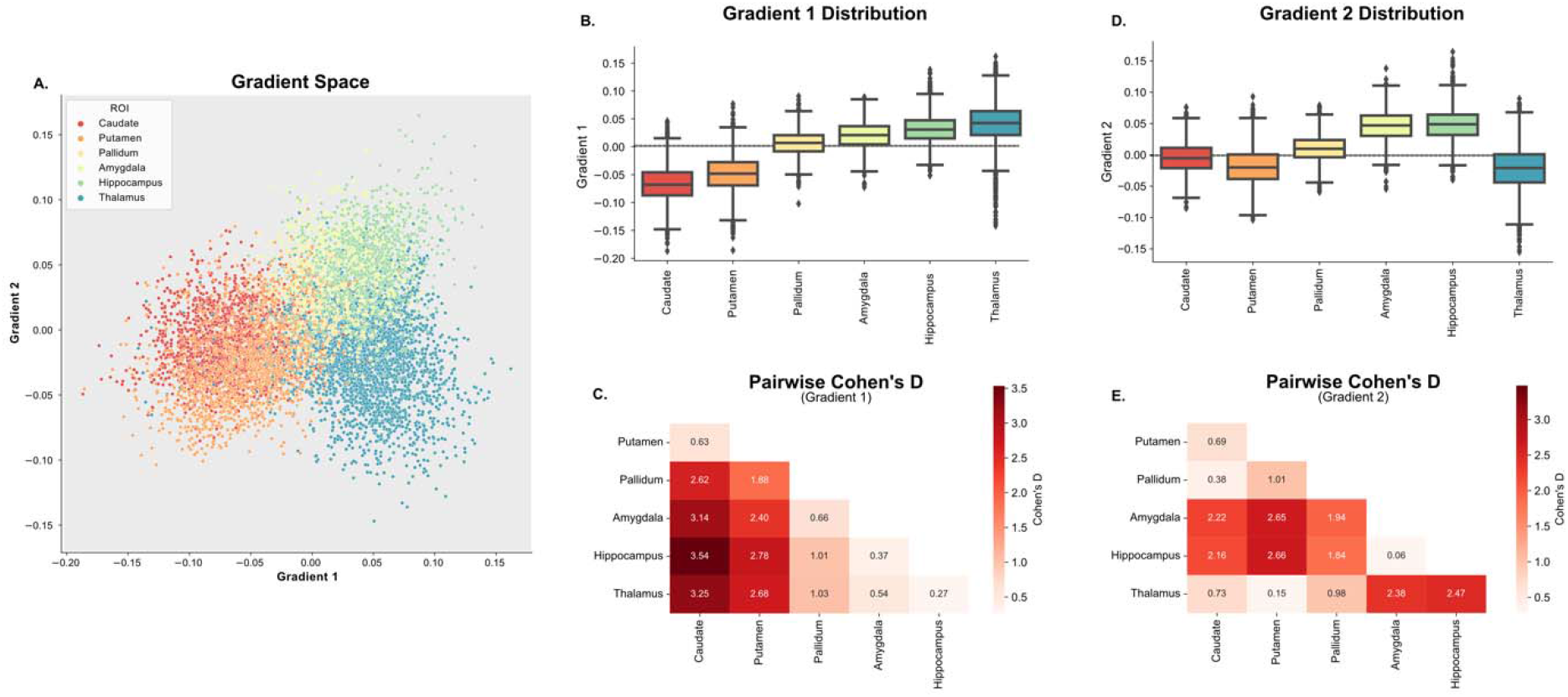
Overview of the Subcortical Functional Gradients: **A**. Average gradient space generated by principal gradient 1 and 2 across all TLE subjects. Different colors represent different subcortical regions of interest (ROIs). Ipsilateral and contralateral structures are assigned the same color in this representation. **B**.,**C**. Boxplots representing the distribution across ROIs for gradient 1 (**B**.) and gradient 2 (**C**.). **D**., **E**. Pairwise Cohen’s D values between each ROI for gradient 1 (**D**.) and gradient 2 (**E**.). Differences between all ROIs were statistically significant (p_BON_ < 0.05)

Mean values across gradient 1 suggest an organization that progresses from regions with lower computational hierarchy (with neural signaling that primarily projects to the thalamus) to regions with higher computational hierarchy (hippocampus and thalamus) (Figure 3B). This is similar to Margulies et. al.’s work in cortical functional gradients, which demonstrated a unimodal to transmodal organization along the first principal gradient ^33^. These results were also replicated in the control cohort (Supplementary Figure 2). All mean ROI pairwise differences across gradient 1 were significant (p<0.001, Bonferroni corrected). However, pairwise Cohen’s D effect sizes between ROIs (Figure 3C) were smallest between the caudate and putamen, as well as between the thalamus, hippocampus and amygdala. The proximity along the dimension of gradient 1 for the hippocampus and amygdala relative to the thalamus was countered by separation along the dimension of gradient 2 (Figure 3D,E), suggesting that gradient 2 represents a separation between functions within lower and higher computational hierarchy domains (e.g. the thalamus is functionally different from the hippocampus and the amygdala, but they are both of high computational hierarchy).

### Subcortical Principal Gradient 1 is Expanded in Temporal Lobe Epilepsy

Previous studies in epilepsy have demonstrated an expansion of the cortical functional gradient 1 in generalized epilepsy relative to controls ^34^. Therefore, we tested whether the subcortical functional gradient was expanded in focal epilepsy relative to controls. We quantified the extent of gradient expansion and contraction by measuring the subject-level variance in the principal gradient across all subcortical ROIs. For R-TLE and L-TLE combined, we found a statistically significant gradient 1 expansion relative to controls (p=0.0180, one-tailed t-test; Cohen’s D=0.54) (Figure 4A). For R-TLE and L-TLE separately, we found a statistically significant expansion of gradient 1 in R-TLE relative to controls (p=0.048, one-tailed t-test, Bonferroni corrected; Cohen’s D=0.68), and a non-statistically significant expansion in L-TLE relative to controls (p=0.138, one-tailed t-test, Bonferroni corrected; Cohen’s D=0.47) (Figure 4B). Figure 4C-E shows representative examples of the spread of gradient 1 across different subjects and different groups. The group level gradient 1 distribution for each group is shown in Supplementary Figure 2. No differences were found for gradient 2 global variance across groups. These findings suggest a subcortical functional principal gradient expansion in focal TLE that is similar to that observed in the cortex and thalamus in generalized epilepsy^34^.

**Figure 4.**
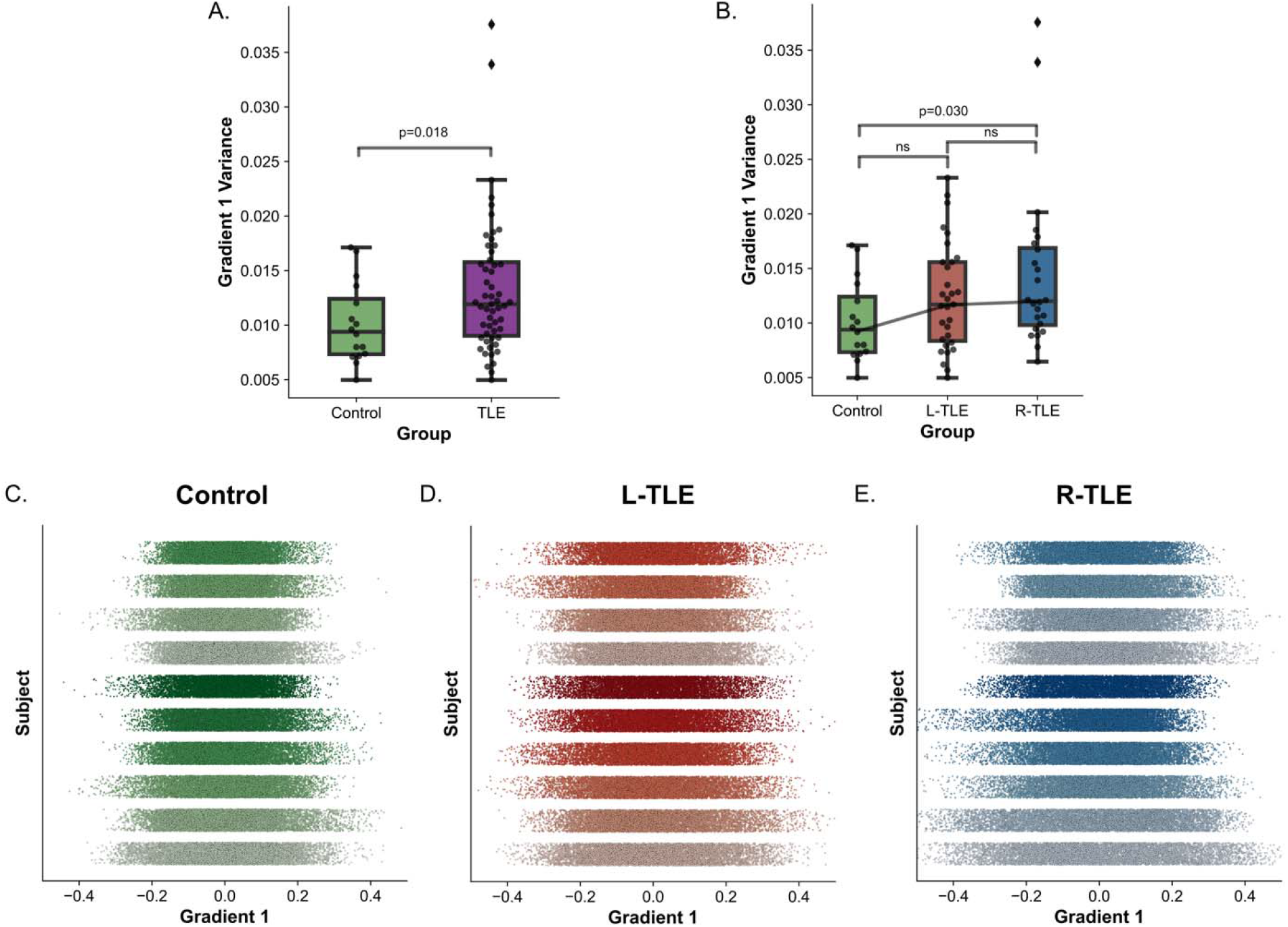
Increased Gradient 1 Global Variance in TLE: Boxplot for global gradient 1 variance across **A**. control and all TLE subjects, and **B**. control, L-TLE, and R-TLE. **C-D**. 1-dimensional scatter of gradient 1 across all ROIs for 10 subjects (one per line) sorted from most gradient 1 variance (bottom) to least (top) for **C**. control, **D**. L-TLE and **E**. R-TLE.

### Hippocampal principal gradient 1 ipsilateral to the SOZ is different in left and right TLE

Given the observed differences in gradient expansion present between left and right TLE, we then explored how the gradient expansion and magnitude differed across subcortical ROIs between left and right TLE. We performed this comparison in two ways: in average gradient space, and in subject space. For the average gradient space comparison, a voxel-wise average was taken across all subjects in each group. This resulted in two average gradient distributions (one for L-TLE and one for R-TLE) for each ROI. For the subject space comparison, the mean and variance of each gradient distribution was computed for each individual subject, and the distribution of subject level gradient mean and variance was compared across groups. The subject space distributions were z-scored relative to the distribution of controls across both left and right ROIs. The results for this analysis, applied to gradient 1 across ROIs ipsilateral to the SOZ, are shown in Figure 5.

**Figure 5.**
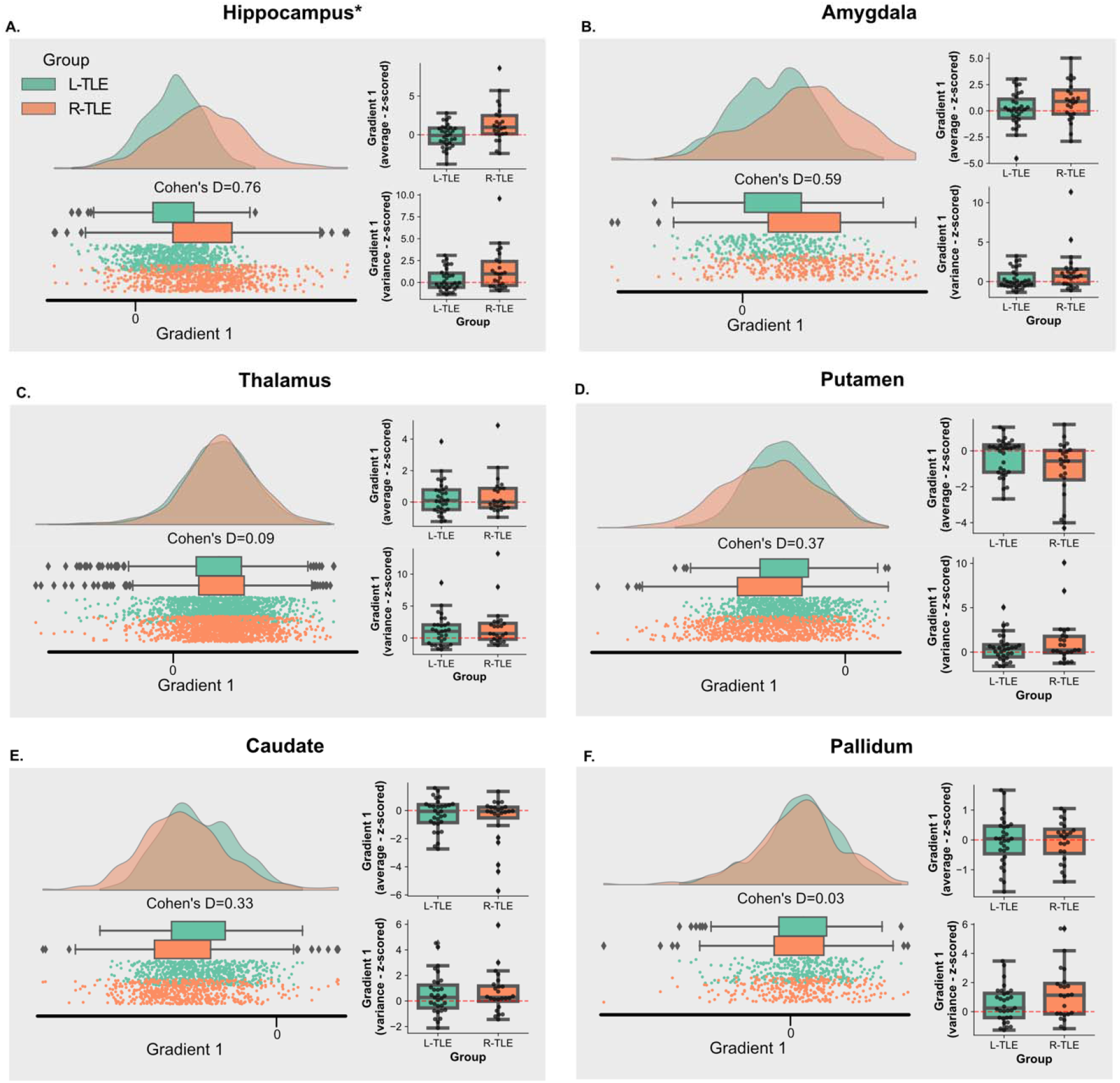
Principal Gradient 1 Across Ipsilateral Subcortical ROIs: **A-F**. Each panel represents a different subcortical ROI, and they show both, the average distribution in gradient space for gradient 1 across subjects in each group (left), and the distribution of individual gradient 1 mean and variance for subjects in each group (right). The individual subject mean and variance were z-scored relative to the distribution of gradient 1 mean and variance for controls in the same ROI, but across bilateral regions.

Average gradient space demonstrates a larger magnitude for the ipsilateral hippocampus in R-TLE relative to L-TLE that trended towards significance (p_BON,PERM_ =0.063; Cohen’s D=0.72). At the subject level, the mean gradient 1 value in the ipsilateral hippocampus was also higher for R-TLE than L-TLE (p_BON,PERM_ =0.032; Cohen’s D=0.87), with the subject level variance also being larger for R-TLE, but not statistically significant (p_BON,PERM_ =0.39; Cohen’s D=0.52). A trend in the same direction was observed in the ipsilateral amygdala: higher group level gradient for R-TLE (p_BON,PERM_ =0.48; Cohen’s D=0.59), and higher subject level gradient mean (p_BON,PERM_ =0.56; Cohen’s D=0.43) and variance (p_BON,PERM_ =0.42; Cohen’s D=0.48), but the findings were not statistically significant. For the thalamus, caudate and pallidum, the differences between L-TLE and R-TLE had small effect sizes (Cohen’s D < 0.50) and no differences were statistically significant (p_BON,PERM_ >0.05). However, a trend in the opposite direction was observed in the putamen and caudate, with a lower group and subject level mean gradient values in R-TLE relative to L-TLE. These findings suggest that the increased global variance in the principal gradient of R-TLE relative to L-TLE and controls, is driven by more extreme (more positive) principal gradient 1 values in the ipsilateral hippocampus. However, potential contributions by more extreme values in other more computationally complex subcortical areas such as the amygdala, and more extreme values in the opposite direction (more negative values) in less computationally complex subcortical areas, like the putamen and caudate, cannot be fully excluded. No sizeable effects nor significant differences were observed for the contralateral ROIs (Supplementary Figure 3).

Using the Bhattacharyya distance, we wanted to validate whether the findings of the 1^st^ principal gradient in the hippocampus would extend to the group level 2-dimensional distribution generated by principal gradients 1 and 2. We found that the Battacharyya distance was statistically significant between the ipsilateral hippocampus of L-TLE and R-TLE (p_PERM_ =0.029, Battacharyya distance=0.10). Distances across other ipsilateral and contralateral ROIs are show in Supplementary Figures 4 and 5.

Overall, these findings demonstrate differences in the ipsilateral hippocampal functional gradient in L-TLE compared to R-TLE, suggesting a cortical connectivity profile in the ipsilateral hippocampus that depends on the laterality of the SOZ.

### Clinical variables other than epilepsy laterality had little association with subcortical functional gradients

We used a linear model to assess whether clinical variables, other than disease laterality, influenced the mean and variance of gradient 1. We found that, for the ipsilateral hippocampus, disease laterality remained the only covariate with a statistically significant association with the gradient 1 mean (p=0.012, corrected across 4 covariates), consistent with the findings in the previous sections of the paper. We also found that the mean gradient 1 of the contralateral caudate had a statistically significant association with the presence of MTS (p=0.012, corrected across 4 covariates). We additionally found for MTS status, negative coefficients for higher computational regions (hippocampus, amygdala, and thalamus), and positive coefficients for lower computational regions (putamen and caudate). This suggests a trend towards gradient 1 contraction in the presence of MTS, or conversely, an expansion in the absence of MTS. Our data further supports this interpretation with a negative coefficient for MTS in a model predicting the global gradient 1 variance, which demonstrates a decrease in the latter quantity (contraction) in the presence of MTS (Supplementary Table 3). We found no other significant associations between the mean, or variance, of gradient 1 and the presence of mesial temporal sclerosis, history of focal to bilateral tonic-clonic seizures, or disease duration across ROIs (Supplementary Table 1 and 2).

### Subcortical functional connectivity gradients were stable across different gradient estimation approaches

Results for the correlation between the gradients across all ROIs computed using different methodologies are shown in Figure 6. Across the different gradient estimation approaches, we found a large correlation across methodologies for gradient 1, with the lowest correlation between approaches that used Laplacian embedding as the dimensionality reduction technique, and approaches that did not (Figure 6C). For gradient 2 (Figure 6D) we found a similar pattern, with an even lower correlation between approaches that used Laplacian embedding as the dimensionality reduction technique, and approaches that did not. We also repeated the Bhattacharyya distance analysis within the ipsilateral hippocampus across all methodologies, and the findings remained statistically significant, with consistent distances, for across all approaches that used a diffusion mapping dimensionality reduction. For Laplacian embedding dimensionality reduction, cosine similarity, Pearson correlation, and Spearman correlation similarities did not produce significant findings, and had very low distances. Finally, for PCA dimensionality reduction, Gaussian kernel and Spearman similarity also had low distances that were not significant. These findings are summarized in Supplementary Table 4.

**Figure 6.**
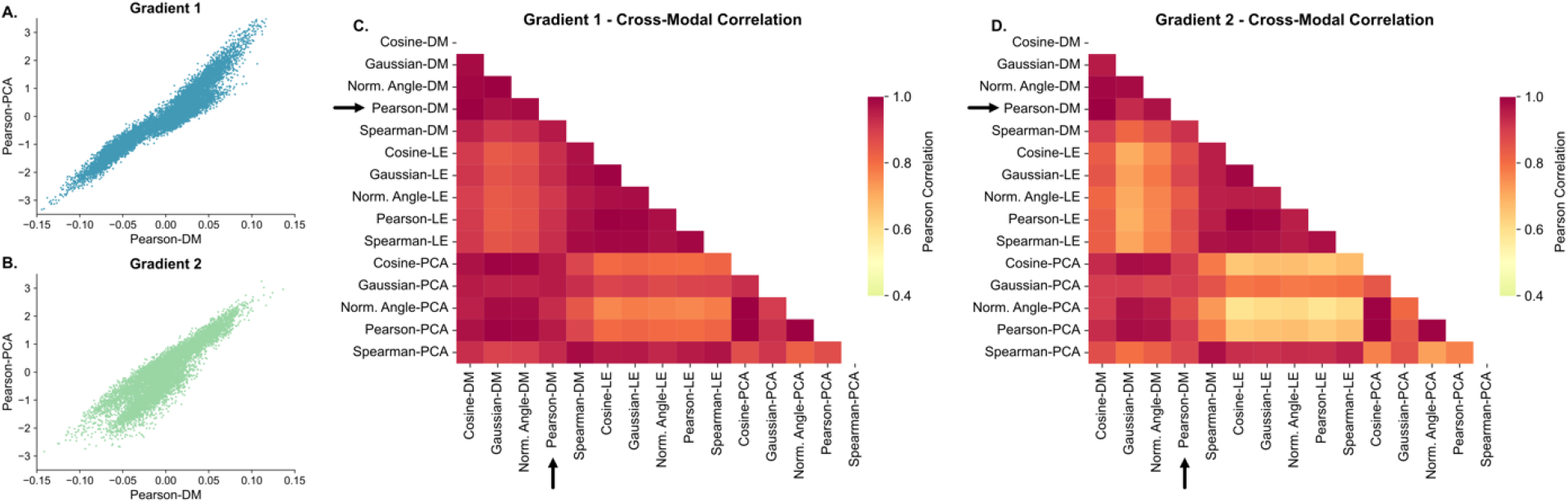
Gradient Stability Across Estimation Methods: **A**. Scatterplot between gradient 1 estimated with a Pearson correlation similarity matrix and diffusion mapping dimensionality reduction (the original approach used in this study), and gradient 1 estimated with a Pearson correlation similarity matrix and principal component analysis dimensionality (PCA) dimensionality reduction. **B**. Same as **A**. but with gradient 2. **C-D**. Absolute value of the Pearson correlation between gradient 1 (**C**.) and gradient 2 (**D**.) for different methods of estimating the similarity matrix and the subsequent dimensionality reduction. Arrows point to the rows and columns corresponding to the Pearson-DM approach used in the main findings of this study. DM – Diffusion mapping, LE – Laplacian embedding, PCA – Principal component analysis.

## Discussion

In this study we describe the subcortical-to-cortical functional connectivity signature of temporal lobe epilepsy via the use of functional connectivity gradients. While previous functional gradient studies in epilepsy have largely focused on cortical gradients, here we provide an overview of subcortical-to-cortical functional gradients, which further supports the involvement of these structures in the pathophysiology of temporal lobe epilepsy. Our results demonstrate a lower to higher computational hierarchy in the subcortex that goes from the putamen to the thalamus along principal gradient 1. We also show an expanded subcortical principal gradient in individuals with TLE subjects relative to healthy controls, which is consistent with previous findings in the cortical gradients of patients with generalized epilepsy^34^, but contradicts previous findings with similar methods in the cortex of patients with TLE^22^. Finally, we show that the gradient expansion in TLE was more pronounced in R-TLE than in L-TLE, with this difference being driven by a larger (more positive) principal gradient 1 value in the ipsilateral hippocampus of R-TLE patients.

A contraction of functional connectivity gradients, relative to neurotypical controls, has been previously demonstrated in several neuropsychiatric disorders, including autism spectrum disorder and schizophrenia^21,35^. In epilepsy, however, studies have demonstrated principal cortical gradient expansion in both generalized epilepsy^34^, and more recently, in newly diagnosed focal epilepsy^36^. Our findings add to the growing body of evidence of an expanded functional gradient in epilepsy, by demonstrating that functional gradient expansion also occurs in the subcortex of patients with TLE. These findings in combination suggest that principal gradient expansion is a characteristic feature of the functional connectome of epilepsy both at the cortical level as well as within the subcortex.

An expanded subcortical functional gradient is evidence of a connectivity profile within subcortical voxels that is more variable in TLE than it is healthy controls. Broadly, this means that two voxels that have a similar connectivity pattern in healthy controls and, therefore, are close together in diffusion embedding space in these, are less likely to have a similar connectivity pattern in individuals with epilepsy, therefore they will be further apart in diffusion embedding space. Furthermore, our findings suggest that the increased variation in connectivity is driven by changes in the hippocampus ipsilateral to the SOZ. Mechanistically, this is consistent with current evidence on TLE, in that we would expect a disrupted connectivity pattern in the hippocampus based on prior structural and functional MRI studies^37–40^. Additionally, some research avenues place the hippocampus as a node within the default mode network^41^, which is where changes have been previously demonstrated to drive gradient expansion in epilepsy^34^. Our findings therefore are consistent and complementary to current neuroimaging research in epilepsy.

Differences between left and right TLE have been demonstrated in the past both in functional and structural neuroimaging studies, and while there is not universal agreement in the literature, many studies report that right TLE has either stronger, or more widespread, abnormalities^3,4,6,42,43^. These reported functional and structural alterations in R-TLE are consistent with the findings of this study, since a stronger expansion of the principal gradient in R-TLE could represent a more widely distributed subcortical, and more specifically, hippocampal, connectivity network. Our findings contribute to the hypothesis of left and right TLE as two distinct epilepsy phenotypes that differ on more than the laterality of the disease, with widespread hippocampo-cortical connectivity abnormalities as part of this phenotype.

In this study, we found that disease laterality was the main factor that seemed to influence gradient values. We also found an association between the contralateral caudate and MTS, which at a larger scale, represented a trend towards global gradient contraction in the presence of MTS, or consequently, an expansion in non-lesional TLE. This might be evidence of a more distributed epileptic network in non-lesional TLE, but our results are preliminary, and further confirmation is needed. As for other clinical factors, history of focal to bilateral tonic-clonic seizures and disease duration were not related to gradient values. There are several reasons as to why this might be the case. First, it is possible that during the dimensionality reduction implemented during gradient generation, the variance of disease laterality dominates over the variance of the other covariates, causing the gradient representation to encode differences in disease laterality much better than differences in other disease factors. Second, it is possible that with more granular ROIs we could obtain more specific gradient changes due to disease factors. For example, quantifying the gradient properties within thalamic subnuclei, such as the mediodorsal nucleus, we might be able to capture differences between patients with and without a history of BTCS^44^. Third, it is possible that the temporal signal-to-noise ratio (tSNR) of the BOLD signal in subcortical structures measured at 3T is not sufficient to discriminate nuances in disease factors at a gradient level, making a higher field strength (e.g. 7T) functional acquisition a more appropriate approach for these questions. Finally, it is also possible that the functional signature of subcortical structures is not sensitive to these disease factors and is instead inherent to the disease itself. This can be particularly true for disease duration, where even in clear structural abnormalities like hippocampal sclerosis, it is still controversial whether disease duration has an impact on hippocampal volume^45–47^.

Our study has several limitations. First, our sample size is moderate. Future multicenter epilepsy studies should attempt to validate our findings using larger sample sizes. Second, gradient representations provide a global characterization of the connectivity from a subcortical voxel to the cortex, however, there is no established methodology for identifying the cortical regions that are driving the subcortical gradient differences. Future work in the functional gradient literature should focus on developing efficient strategies for mapping the gradient differences back to target cortical voxels. This would allow researchers in the field to confirm whether the differences between R and L-TLE that we reported in the ipsilateral hippocampus are driven by connectivity differences in the default mode network, or other cortical regions. Finally, we made use of subcortical ROIs that were defined a priori through a standard atlas. While appropriate for this exploratory study, future studies should leverage subject-specific subcortical segmentations, such as those provided by FreeSurfer^48^, ASHS^49^ and THOMAS^50^. Additionally, usage of ultra-high field imaging at 7T can not only help get more accurate segmentations of these structures, but also improved tSNR in the BOLD signal, which would precisely localize signals within the subcortex, further elucidating differences between epilepsy subtypes and disease factors in gradient space.

## Conclusions

In this study we describe the subcortical-to-cortical functional connectivity signature of temporal lobe epilepsy through functional connectivity gradients, demonstrating an expansion of the principal subcortical gradient in individuals with epilepsy relative to healthy controls. These findings indicate gradient expansion as a functional connectome phenotype in epilepsy. We also demonstrate differences in the gradient between L-TLE and R-TLE that may be driven by changes in the hippocampus ipsilateral to the seizure onset zone.

## Data Availability

All data produced in the study will be made publicly available once it is published.

## Data Availability Statement

The processed data and methodological pipeline used in this study will be made available on GitHub upon publication of the manuscript. The repository will include all necessary files and documentation to reproduce the results presented in the paper. The repository will be made publicly accessible to ensure that the data and methods used in this study can be transparently and independently verified.

## Guidelines on Research Ethics and Publishing

We confirm that we have read the Journal’s position on issues involved in ethical publication and affirm that this report is consistent with those guidelines.

