## Supplement for "Subcortical Functional Connectivity Gradients in Temporal Lobe Epilepsy"

Supplementary Figures and Tables

**Supplementary Table 1 –** Regression coefficients and corresponding p-values for linear model of disease factors predicting the z-scored mean of gradient 1 at each subcortical ROI

**I-Hippocampus**

| | Laterality | MTS | BTCS | Duration |
|---|---|---|---|---|
| **β** | **-0.0197** | -0.0032 | -0.0035 | -0.0003 |
| **p-value** | **0.004** | 0.665 | 0.631 | 0.332 |

**C-Hippocampus**

| | Laterality | MTS | BTCS | Duration |
|---|---|---|---|---|
| **β** | -0.0082 | 0.0004 | 0.0014 | -0.0002 |
| **p-value** | 0.151 | 0.955 | 0.828 | 0.323 |

**I-Amygdala**

| | Laterality | MTS | BTCS | Duration |
|---|---|---|---|---|
| **β** | -0.0133 | 0.0027 | -0.0002 | 0.0002 |
| **p-value** | 0.164 | 0.799 | 0.983 | 0.655 |

**C-Amygdala**

| | Laterality | MTS | BTCS | Duration |
|---|---|---|---|---|
| **β** | -0.0036 | 0.0107 | 0.002 | 0.0002 |
| **p-value** | 0.637 | 0.206 | 0.811 | 0.554 |

**I-Thalamus**

| | Laterality | MTS | BTCS | Duration |
|---|---|---|---|---|
| **β** | -0.0041 | -0.0025 | -0.0122 | -0.0005 |
| **p-value** | 0.58 | 0.757 | 0.135 | 0.121 |

**C-Thalamus**

| | Laterality | MTS | BTCS | Duration |
|---|---|---|---|---|
| **β** | 0.0041 | 0.0004 | -0.0078 | -0.0003 |
| **p-value** | 0.626 | 0.968 | 0.392 | 0.39 |

**I-Caudate**

| | Laterality | MTS | BTCS | Duration |
|---|---|---|---|---|
| **β** | 0.0096 | -0.0034 | **0.029** | 0.0006 |
| **p-value** | 0.351 | 0.765 | **0.012** | 0.13 |

**C-Caudate**

| | Laterality | MTS | BTCS | Duration |
|---|---|---|---|---|
| **β** | 0.0046 | 0.0005 | 0.0159 | 0.0008 |
| **p-value** | 0.652 | 0.962 | 0.161 | 0.071 |

**I-Putamen**

| | Laterality | MTS | BTCS | Duration |
|---|---|---|---|---|
| **β** | 0.0122 | -0.0003 | 0.0058 | 0.0004 |
| **p-value** | 0.047 | 0.96 | 0.377 | 0.119 |

**C-Putamen**

| | Laterality | MTS | BTCS | Duration |
|---|---|---|---|---|
| **β** | 0.0067 | 0.0003 | 0.0005 | 0.0002 |
| **p-value** | 0.371 | 0.972 | 0.953 | 0.567 |

**I-Pallidum**

| | Laterality | MTS | BTCS | Duration |
|---|---|---|---|---|
| **β** | 0.002 | 0.0076 | -0.0083 | -0.0002 |
| **p-value** | 0.674 | 0.152 | 0.113 | 0.39 |

**C-Pallidum**

| | Laterality | MTS | BTCS | Duration |
|---|---|---|---|---|
| **β** | -0.0063 | 0.0021 | -0.0089 | -0.0001 |
| **p-value** | 0.423 | 0.81 | 0.299 | 0.697 |

**Supplementary Table 2 –** Regression coefficients and corresponding p-values for linear models of disease factors predicting the z-scored variance of gradient 1 at each subcortical ROI

**I-Hippocampus**

| | Laterality | MTS | BTCS | Duration |
|---|---|---|---|---|
| **β** | **-0.0034** | -0.0027 | -0.0005 | -4.04E-05 |
| **p-value** | **0.01** | 0.058 | 0.705 | 0.435 |

**C-Hippocampus**

| | Laterality | MTS | BTCS | Duration |
|---|---|---|---|---|
| **β** | -0.0027 | -0.0024 | 0.0004 | -7.75E-05 |
| **p-value** | 0.041 | 0.108 | 0.789 | 0.146 |

**I-Amygdala**

| | Laterality | MTS | BTCS | Duration |
|---|---|---|---|---|
| **β** | -0.0033 | -0.0026 | -0.0024 | -0.0001 |
| **p-value** | 0.034 | 0.132 | 0.152 | 0.087 |

**C-Amygdala**

| | Laterality | MTS | BTCS | Duration |
|---|---|---|---|---|
| **β** | -0.0025 | -0.0019 | -0.0005 | -8.04E-05 |
| **p-value** | 0.063 | 0.211 | 0.757 | 0.137 |

**I-Thalamus**

| | Laterality | MTS | BTCS | Duration |
|---|---|---|---|---|
| **β** | -0.0032 | -0.0028 | 0.0013 | -7.32E-05 |
| **p-value** | 0.029 | 0.088 | 0.431 | 0.213 |

**C-Thalamus**

| | Laterality | MTS | BTCS | Duration |
|---|---|---|---|---|
| **β** | -0.0024 | -0.0028 | 0.0011 | -8.64E-05 |
| **p-value** | 0.151 | 0.126 | 0.554 | 0.199 |

**I-Caudate**

| | Laterality | MTS | BTCS | Duration |
|---|---|---|---|---|
| **β** | -0.001 | -0.0013 | 6.35E-05 | -2.33E-05 |
| **p-value** | 0.397 | 0.333 | 0.961 | 0.627 |

**C-Caudate**

| | Laterality | MTS | BTCS | Duration |
|---|---|---|---|---|
| **β** | -0.0015 | -0.0014 | 0.0006 | -2.59E-05 |
| **p-value** | 0.189 | 0.255 | 0.63 | 0.571 |

**I-Putamen**

| | Laterality | MTS | BTCS | Duration |
|---|---|---|---|---|
| **β** | -0.002 | -0.0014 | 0.0005 | -2.81E-05 |
| **p-value** | 0.161 | 0.379 | 0.738 | 0.623 |

**C-Putamen**

| | Laterality | MTS | BTCS | Duration |
|---|---|---|---|---|
| **β** | -0.0018 | -0.0015 | 0.0001 | -7.69E-05 |
| **p-value** | 0.175 | 0.316 | 0.929 | 0.161 |

**I-Pallidum**

| | Laterality | MTS | BTCS | Duration |
|---|---|---|---|---|
| **β** | -0.002 | -0.0019 | -0.0003 | -3.23E-05 |
| **p-value** | 0.055 | 0.093 | 0.783 | 0.428 |

**C-Pallidum**

| | Laterality | MTS | BTCS | Duration |
|---|---|---|---|---|
| **β** | -0.0032 | -0.0031 | 0.0015 | -8.03E-05 |
| **p-value** | 0.033 | 0.061 | 0.347 | 0.184 |

**Supplementary Table 3 -** Regression coefficients and corresponding p-values for linear models of disease factors predicting the global variance of gradient 1 across all subcortical ROIs

| | Global Variance | | | |
|---|---|---|---|---|
| | **Laterality** | **MTS** | **BTCS** | **Duration** |
| **β** | -0.8413 | -0.9904 | 0.3233 | -0.0155 |
| **p-value** | 0.086 | 0.069 | 0.542 | 0.427 |

**Supplementary Table 4 -** Bhattacharyya Distance and corresponding p-value between the 2D distribution generated by gradient 1 and gradient 2 in L-TLE and R-TLE subjects in the ipsilateral hippocampus, computed using different similarity metrics and dimensionality reduction approaches for gradient estimation. DM: Diffusion mapping. LE: Laplacian embedding. PCA: principal component analysis.

| Method | Bhattacharyya Distance | p-value |
|---|---|---|
| **Cosine-DM** | **0.107963972** | **0.010989011** |
| **Gaussian-DM** | **0.095780277** | **0.020979021** |
| **Norm. Angle-DM** | **0.106681173** | **0.01998002** |
| **Pearson-DM** | **0.097407341** | **0.018981019** |
| **Spearman-DM** | **0.065434815** | **0.03996004** |
| Cosine-LE | 0.02770632 | 0.087912088 |
| **Gaussian-LE** | **0.034955792** | **0.042957043** |
| **Norm. Angle-LE** | **0.044388407** | **0.047952048** |
| Pearson-LE | 0.025584961 | 0.116883117 |
| Spearman-LE | 0.022037481 | 0.084915085 |
| **Cosine-PCA** | **0.1069308** | **0.022977023** |
| Gaussian-PCA | 0.059444441 | 0.06993007 |
| **Norm. Angle-PCA** | **0.111091021** | **0.016983017** |
| **Pearson-PCA** | **0.10620882** | **0.016983017** |
| Spearman-PCA | 0.031851973 | 0.228771229 |

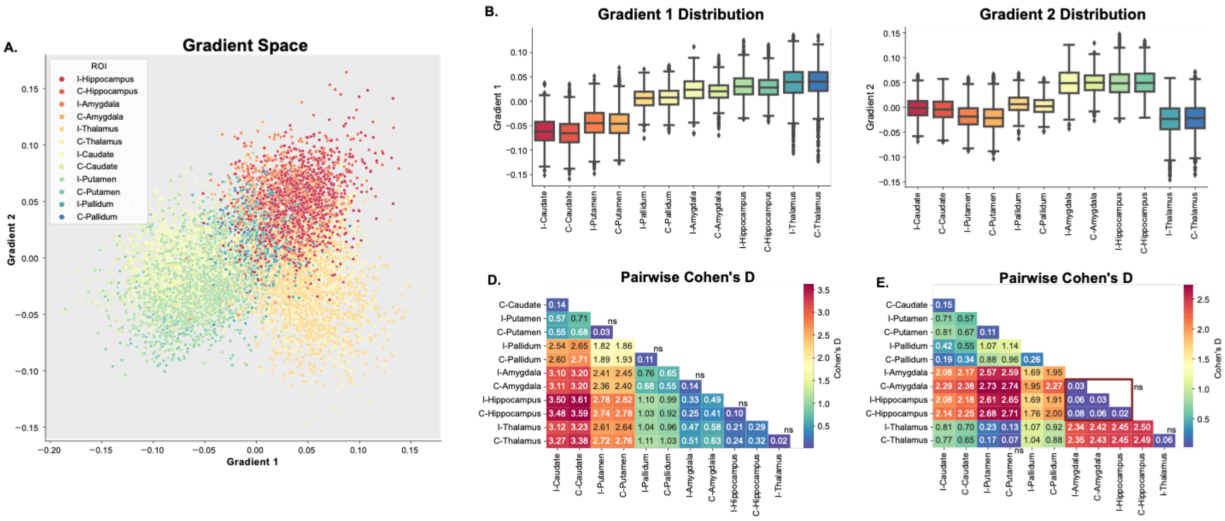


**Supplementary Figure 1 - Overview of the Subcortical Functional Gradient Across Ipsilateral and Contralateral ROIs: A.** Average gradient space generated by principal gradient 1 and 2 across all TLE subjects. Different colors represent different subcortical regions of interest (ROIs). **B.,C.** Boxplots representing the distribution across ROIs for gradient 1 (**B.**) and gradient 2 (**C.**). **D.**, **E.** Pairwise Cohen's D values between each ROI for gradient 1 (**D.**) and gradient 2 (**E.**). Differences between ROIs were statistically significant ($p_{FDR} < 0.05$) unless specified otherwise (n.s.).

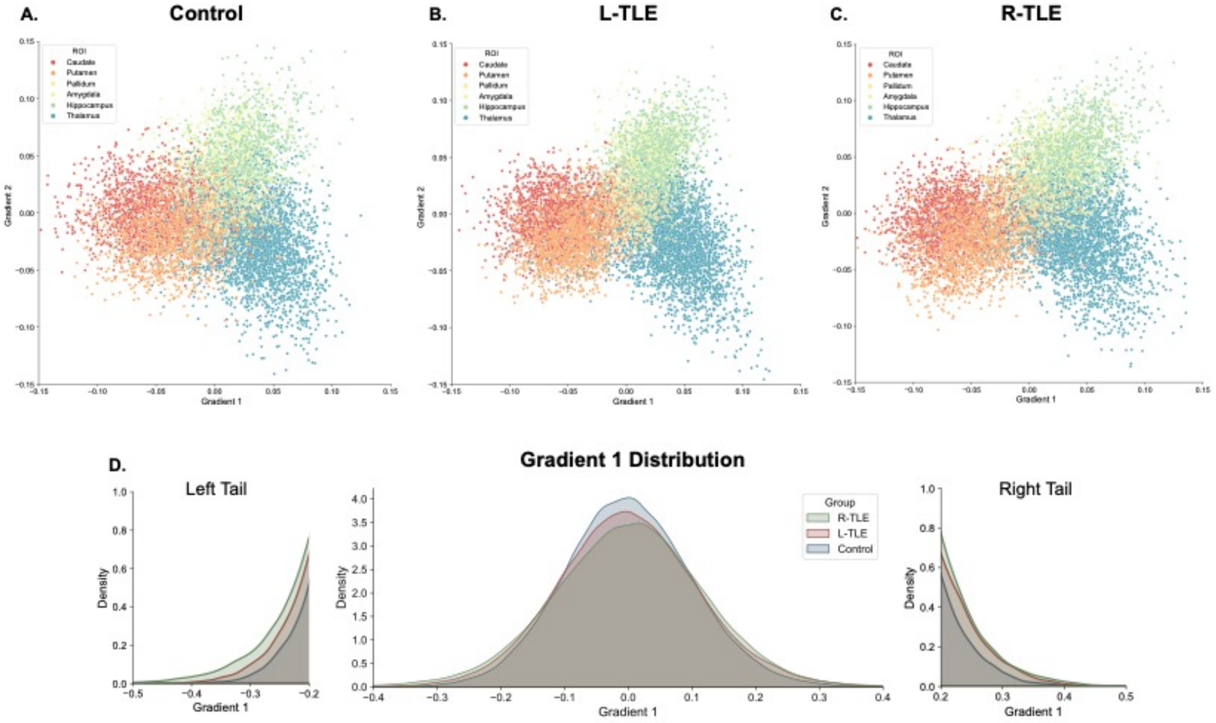


**Supplementary Figure 2 - Overview of the Subcortical Functional Gradient Across ROIs in Healthy Controls, L-TLE and R-TLE: A-C.** Average gradient space generated by principal gradient 1 and 2 across all **A.** control subjects, **B.** L-TLE subjects and **C.** R-TLE subjects. Ipsilateral and contralateral structures are assigned the same color in this representation. **D.** Distribution of Gradient 1 across all subjects and ROIs for each subgroup. The left and right tail show an expansion of gradient 1 for left and right TLE relative to controls.

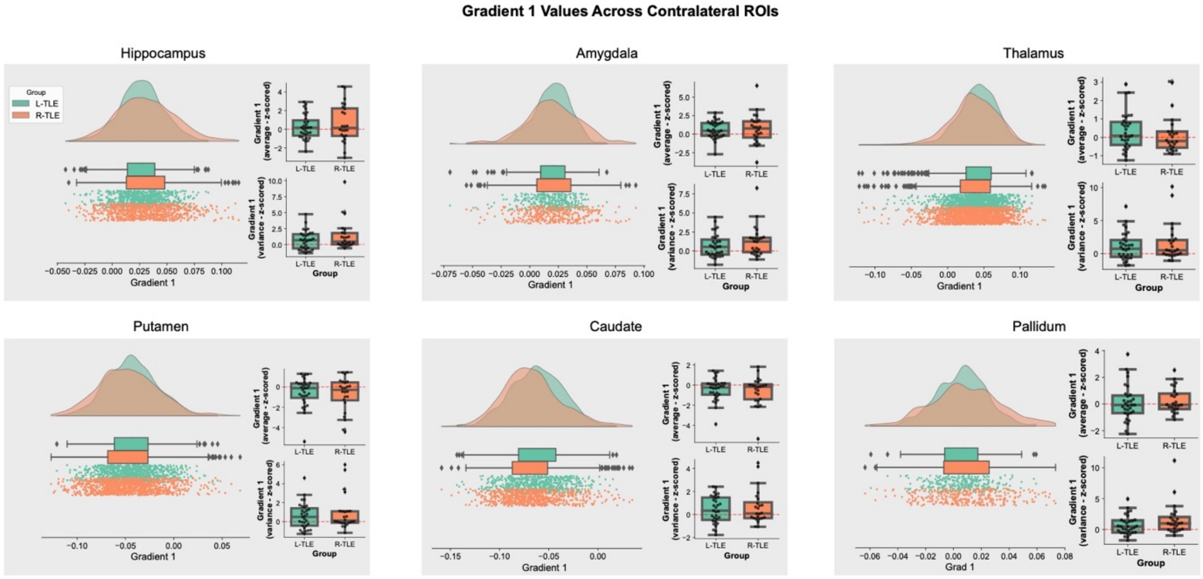


**Supplementary Figure 3 - Principal Gradient 1 Across Contralateral Subcortical ROIs: A-F.** Each panel represents a different subcortical ROI, and they show both, the average distribution in gradient space for gradient 1 across subjects in each group (left), and the distribution of individual gradient 1 mean and variance for subjects in each group (right). The individual subject mean and variance were z-scored relative to the distribution of gradient 1 mean and variance for controls in the same ROI, but across bilateral regions.

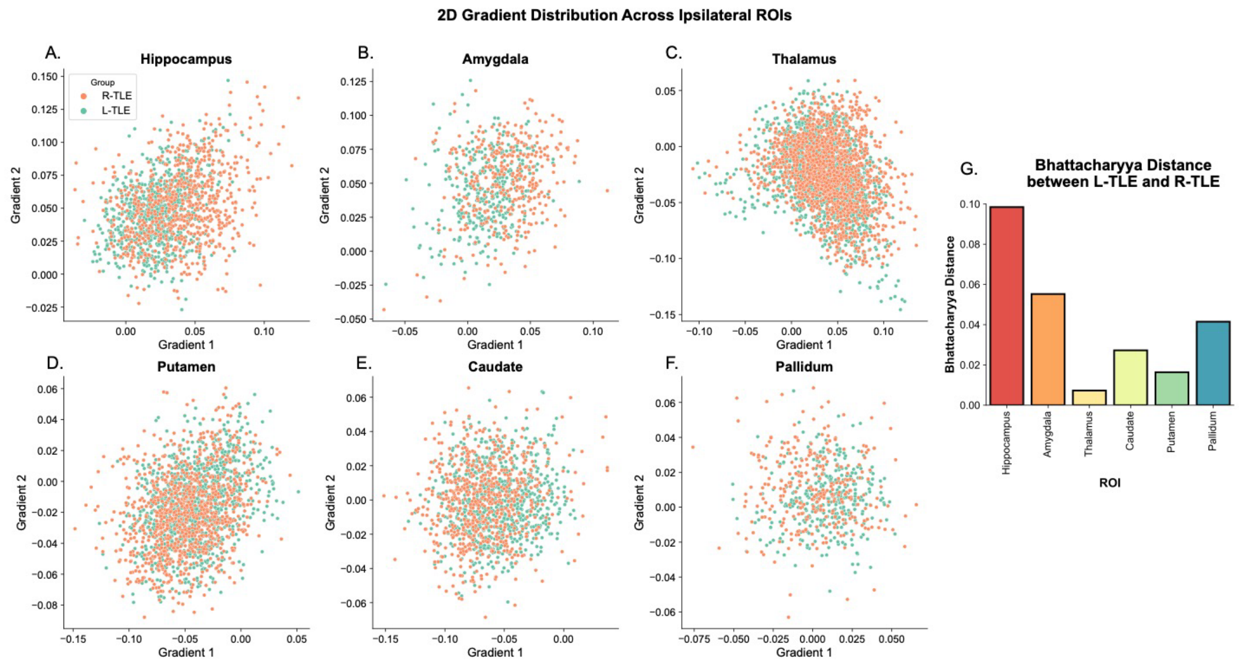


**Supplementary Figure 4 – 2-Dimensional Gradient Distribution Across Ipsilateral ROIs: A-F.** Group average 2-dimensional distribution generated by subcortical functional gradient 1 and 2 of R-TLE and L-TLE across ipsilateral ROIs. **G.** Bhattacharyya distance between the distribution of L-TLE and R-TLE across ROIs.

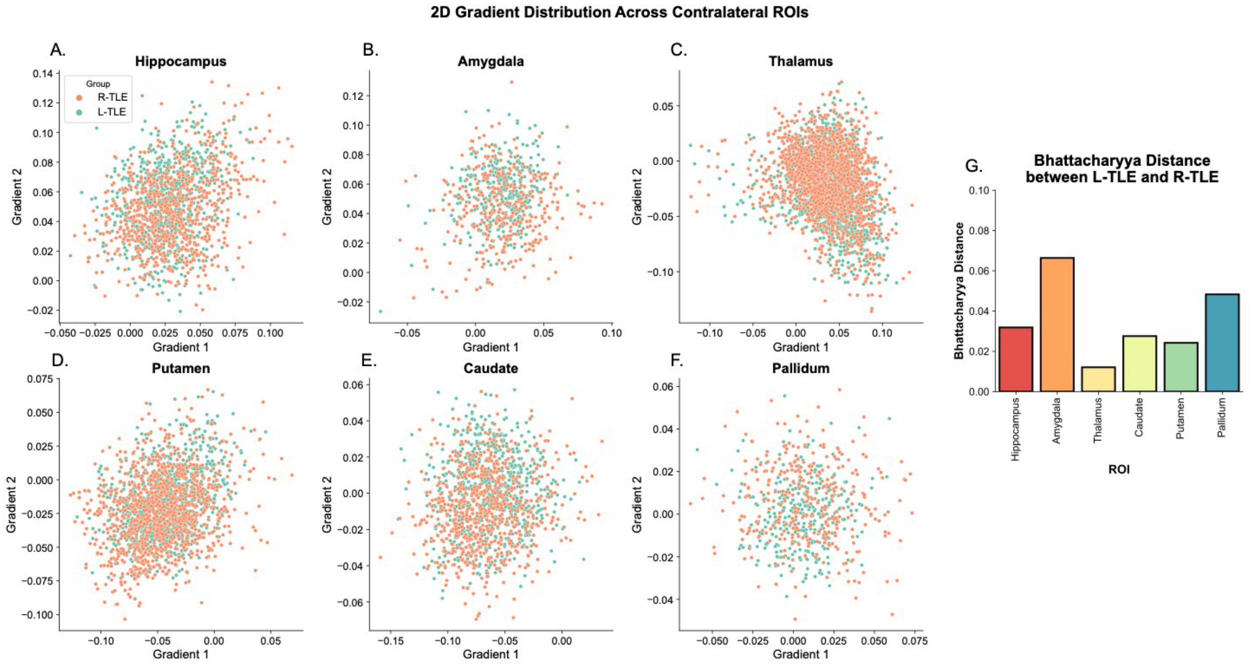


**Supplementary Figure 5 – 2-Dimensional Gradient Distribution Across Contralateral ROIs: A-F.** Group average 2-dimensional distribution generated by subcortical functional gradient 1 and 2 of R-TLE and L-TLE across contralateral ROIs. **G.** Bhattacharyya distance between the distribution of L-TLE and R-TLE across ROIs.
